# Sun protection and skin cancer screening after childhood cancer – a report from the Swiss Childhood Cancer Survivor Study (SCCSS)

**DOI:** 10.1101/2025.11.15.25340311

**Authors:** Carina Nigg, Maša Žarković, Philippa Jörger, Eva Maria E. Tinner, Calogero Mazzara, Eva Brack, Paul Castle, Alexander Navarini, Christina Schindera, Claudia E Kuehni

## Abstract

**Background:** Childhood cancer survivors (CCS) face elevated skin cancer risk, especially after radiotherapy or hematopoietic stem cell transplantation (HSCT). We evaluated the prevalence and predictors of sun protection, sunburn, and physician skin examination (PSE) among CCS in Switzerland.

**Methods:** We surveyed CCS diagnosed <21 years and surviving ≥5 years after diagnosis about sun protection, sunburns during last summer, and PSE within the last year. We retrieved cancer-related data from the Swiss Childhood Cancer Registry and used multivariable logistic regression, stratified by age group, to identify predictors.

**Results:** We included 1,048 children (5-15 years), 572 adolescents (16-19 years), and 1,959 adults (≥20 years). Regular sun protection was reported by 89% of children, 65% of adolescents, and 77% of adults, and sunburns by 23%, 49%, and 43%. PSE prevalence among those treated with radiotherapy was 21%, 18%, and 17%, and among HSCT recipients 36%, 28%, and 28%. Radiotherapy was unrelated to sun protection and PSE, but associated with fewer sunburns (OR=0.63-0.77). HSCT recipients were more likely to have attended a PSE (OR=2.06-3.75), but not radiotherapy recipients. Across age groups, survivors born more recently were less likely to protect from sun (OR range=0.94-0.97) and more likely to report sunburn (OR=1.04-1.14).

**Conclusion:** Survivors protect insufficiently from sun and only few who are particularly at risk for skin cancer due to their treatment history attend PSEs as recommended by the Children’s Oncology Group. Healthcare practitioners should systematically integrate yearly PSE after radiotherapy or HSCT and encourage consistent sun protection, particularly among younger generations and adolescents.

## Introduction

Melanoma skin cancer incidence, the most serious skin cancer type, has increased significantly in Switzerland over the past 40 years: While 4,000 melanomas were registered in Switzerland between 1982 and 1986, , tthis number quadrupled to 16,500 cases between 2017 and 2021.^1^ This makes melanomas the fifth most common cancer type in Switzerland,^2^ and Switzerland one of the leading melanoma incidence countries in Europe.^3^ Genetic predisposition, especially a fair, sun-sensitive skin type, and environmental exposure, particularly ultraviolet radiation through sun exposure, are the main risk factors for developing skin cancer.^4^ For CCS, certain cancer treatments, such as radiation, chemotherapy including alkylating agents or bleomycin, or hematopoietic stem cell transplantation (HSCT) increase the risk for developing basal and squamous cell carcinomas,^5,6^ and melanomas.^7^ CCS are twice as likely to develop melanomas compared to their peers without cancer history,^8^ and have a 30-fold higher risk of developing basal cell carcinoma (BCC) already in young adulthood. The Children’s Oncology Group (COG) recommends that survivors treated with radiation or HSCT have a physician skin examination (PSE) once a year and that all survivors protect from sun^9^ to reduce skin cancer mortality.^10–12^ Yet, we know little about sun protection and PSE after childhood cancer. Existing studies focus primarily on the US,^13–17^ with different sun protection practices compared to Europe,^18^ and on adult CCS. ^13–15,19,20^ Most findings originate from studies conducted 15 to 20 years ago.^13,14,17,19^ We aim to describe sun protection, sunburns, and PSE in participants of the Swiss Childhood Cancer Survivor Study (SCCSS), a population-based study of pediatric, adolescent, and adult CCS with data collected 2007-2022. In particular, this study evaluates prevalence and predictors of sun protection, sunburn, and PSE, and assesses compliance with PSE as recommended by the COG.

## Methods

### Study design and study population

The Swiss Childhood Cancer Survivor Study (SCCSS) is a population-based study nested in the Childhood Cancer Registry (ChCR). Detailed methods of the SCCSS are available elsewhere.^21^ The ChCR includes all children and adolescents diagnosed with leukemias, lymphomas, central nervous system (CNS) tumors, malignant solid tumors, or Langerhans cell histiocytosis prior to age of 20 years in Switzerland since 1976.^22^ In the SCCSS, we sent questionnaires to all CCS who had survived ≥5 years since their initial diagnosis.

Questionnaires are available in German, French, and Italian. For this study, we included CCS who participated in the SCCSS between 2007 and 2022. For CCS aged 5-15 years, we asked their parents to fill in the questionnaire. Ethical approval was granted by the ethics committee of the canton of Bern, Switzerland (KEK-BE: 166/2014 and 2021-01462).

### Sun protection, sunburn, and physician skin examination

#### Sun protection

We asked all participants how consistently they (or their child, for parents) protect themselves from the sun (e.g., using sunscreen, seeking shade, or wearing protective clothing). Response options ranged from “Always” to “(Almost) never” (*Supplementary Table 1*). For risk factor analysis, we defined good sun protection as protecting always or mostly from sun, and poor sun protection as protecting rarely or never from sun.

### Sunburn

We asked all participants whether they (or their child, for parents) had experienced one or more sunburns in the past summer (response options no/yes).

### Physician skin examination

We asked all participants whether they had ever had their skin or moles examined by a physician within the last 12 months, more than 12 months ago, or never. We defined compliance with the COG guidelines as having had the skin examined within the last 12 months, and no compliance if they had never had a PSE or if it was more than 12 months ago.

### Explanatory variables

#### Sociodemographic characteristics

We assessed the following sociodemographic characteristics via questionnaires: Swiss language region, parental and participant nationality, and parental education. We categorized language region into German and French/Italian. For migration background and parental education, we applied the definitions of the Federal Statistics Office.^23,24^ If both parents were born Swiss or if the participant and at least one parent were born Swiss, we defined this as no migration background.^23^ For those with migration background, we applied the United Nations’ geographic coding scheme to obtain some information on their cultural background.^25^ For educational background, we considered the highest educational level achieved by either parent. We divided education into three categories: primary (compulsory schooling only), secondary (higher schooling or vocational training), or tertiary (upper vocational education or university or technical college).^24^

### Cancer-related characteristics

We received the following cancer-related characteristics from the ChCR: sex, date of birth, age at diagnosis, and diagnosis according to the International Classification of Childhood Cancer, third edition (ICCC-3).^26^ Treatment information included chemotherapy, radiotherapy, HSCT, and relapse (all no/yes).

### Skin type

We used the Fitzpatrick scale to assess skin phototyping. This six-level system classifies skin type based on genetic predisposition, skin reaction to sun exposure, and tanning ability.^27^

### Statistical analysis

We stratified analyses by age group: children (5-15 years), adolescents (16-19 years), and adults (>20 years). We included all participants in the analysis who responded to at least one of the questions in the questionnaire section about sun protection and PSE.^9^ We calculated the number and proportion of CCS for each of the response options for sun protection, sunburn, and PSE. To assess compliance with COG recommendations, we further assessed number and proportion of CCS treated with radiotherapy or HSCT who attended a PSE within the last 12 months.^9^ To investigate factors associated with sun protection, sunburn, and PSE, we applied multivariable logistic regression analysis. We decided *a priori* to include age at study, sex, migration background, language region, parental education, radiotherapy, and HSCT as exposures in all models, and also considered year of study and birth year to account for potential age–period–cohort effects. Because age, year of study, and birth year are perfectly collinear, we compared models including these predictors separately and in combination.^28^ Model fit based on Akaike (AIC) and Bayesian (BIC) information criteria indicated that including all three was unnecessary. Models including birth year consistently showed the best fit when also considering all other exposures (*Supplementary Table 2*) and were thus used in final analyses, while we retained models with year of study for sensitivity analyses. As we had only asked a subsample of study participants on skin type, we performed additional sensitivity analyses including this variable. Given that several outcomes were common, we also conducted Poisson regression with robust error variances to estimate the relative risk (RR) and enable direct comparison between exposure groups.^29^

## Results

In total, of 5,940 eligible survivors, 5,427 were contacted. Of those, 3,768 (69%) filled in the questionnaire, and 3,579 (66%) responded to at least one question regarding sun exposure or PSE (1,048 pediatric, 572 adolescent, and 1,959 adult CCS, *Supplementary Figure 1*). Median age at study was 21 years (interquartile range [IQR] 15-27), median age at diagnosis 7 years (IQR 3-13), and median time since diagnosis 12 years (IQR 8-19). Leukemia was the most common cancer diagnosis (32%), followed by lymphomas (17%) and CNS tumors (16%). Overall, 77% had received chemotherapy, 31% radiotherapy, and 7% HSCT (*Table 1*).

**Table 1.** Characteristics of childhood cancer survivors included in sun exposure and physician skin examination study.

|  | Children (5-15 years)<br>N = 1,048 | Adolescents (16-19<br>years)<br>N = 572 | Adults (≥ 20<br>years)<br>N = 1,959 |
| --- | --- | --- | --- |
| **Socio-demographic characteristics** |  |  |  |
| *Age at time of questionnaire, median [IQR]* | 12 [10-14] | 18 [17-19] | 27 [23-33] |
| *Year of birth [IQR]* | 2003 [1999-2007] | 1992 [1991-1994] | 1982 [1976-1988] |
| *Year of study participation* |  |  |  |
| 2007-2013 | 450 (43%) |  |  |
| 2007-2009 |  | 257 (45%) | 1,076 (55%) |
| 2010-2013 |  | 179 (31%) | 413 (21%) |
| 2015-2017 | 314 (30%) | 73 (13%) | 232 (12%) |
| 2021-2022 | 284 (27%) | 63 (11%) | 238 (12%) |
| *Female* | 464 (44%) | 273 (48%) | 937 (48%) |
| *Migration background* | 229 (22%) | 119 (21%) | 341 (17%) |
| *UN Geographic region\** |  |  |  |
| Northern/Western/Eastern Europe | 835 (80%) | 465 (82%) | 1590 (83%) |
| Southern Europe | 118 (11%) | 79 (14%) | 207 (11%) |
| Rest of the world | 45 (4%) | 23 (4%) | 20 (1%) |
| *Language* |  |  |  |
| German | 728 (70%) | 399 (70%) | 1382 (71%) |
| French | 276 (26%) | 155 (27%) | 516 (26%) |
| Italian | 44 (4%) | 18 (3%) | 61 (3%) |
| *Parental highest education\** |  |  |  |
| Primary education | 81 (8%) | 33 (5.8%) | 193 (9.9%) |
| Secondary education | 361 (34%) | 344 (60.1%) | 1001 (51.1%) |
| Tertiary education | 577 (55%) | 171 (29.9%) | 657 (33.5%) |
| **Cancer-related characteristics** |  |  |  |
| *ICCC3 main group* |  |  |  |
| Leukemias | 417 (40%) | 188 (33%) | 548 (28%) |
| Lymphomas | 66 (6%) | 84 (15%) | 466 (24%) |
| CNS tumors | 169 (16%) | 104 (18%) | 291 (15%) |
| Neuroblastoma | 93 (9%) | 31 (5%) | 52 (3%) |
| Retinoblastoma | 61 (6%) | 14 (2%) | 28 (1%) |
| Renal tumor | 84 (8%) | 44 (8%) | 71 (4%) |
| Hepatic tumor | 15 (1%) | 3 (<1%) | 12 (<1%) |
| Malignant bone tumors | 18 (2%) | 19 (3%) | 117 (6%) |
| Soft tissue sarcomas | 65 (6%) | 36 (6%) | 124 (6%) |
| Germ cell tumors | 23 (2%) | 15 (3%) | 127 (7%) |
| Other | 4 (<1%) | 8 (1%) | 68 (4%) |
| Langerhans cell histiocytosis | 33 (3%) | 26 (5%) | 55 (3%) |
| *Age at diagnosis, median [IQR]* | 3 [2-5] | 6 [3-10] | 12 [6-16] |
| *Time since diagnosis, median [IQR]* | 8 [7-10] | 11 [8-15] | 18 [11-23] |
| *Year of diagnosis, median [IQR]* | 2010 [2003-2011] | 2000 [1995-2004] | 1990 [1985-2000] |
| *Radiotherapy* | 197 (19%) | 164 (29%) | 749 (38%) |
| *Chemotherapy* | 859 (82%) | 454 (79%) | 1456 (74%) |
| *HSCT* | 82 (8%) | 38 (7%) | 115 (6%) |
| *Relapse* | 37 (4%) | 15 (3%) | 55 (3%) |

\* Percentages are based upon the available N, not adding up to 100%. UN = United Nations, ICCC3 = International Classification of Childhood Cancer, 3<sup>rd</sup> edition, CNS = Central nervous system, HSCT = Hematopoietic stem cell transplantation

### Prevalence of sun protection, sunburn and PSE

Overall, 79% of survivors reported good sun protection (*Supplementary Table 3*). Sun protection differed by age group (*Figure 1*): 88% of children, 65% of adolescents, and 77% of adults reported protecting always or mostly from sun. Nonetheless, 38% reported at least one sunburn the previous summer, with the highest prevalence among adolescents (49%). PSE compliance with the COG recommendation was low (*Figure 2, Supplementary Table 4*). Among survivors treated with radiotherapy, 17% had a PSE within the past 12 months compared with 15% of those without radiotherapy. Among HSCT survivors, 29% reported a recent PSE versus 15% without HSCT. These patterns were similar across age groups.

**Figure 1.**
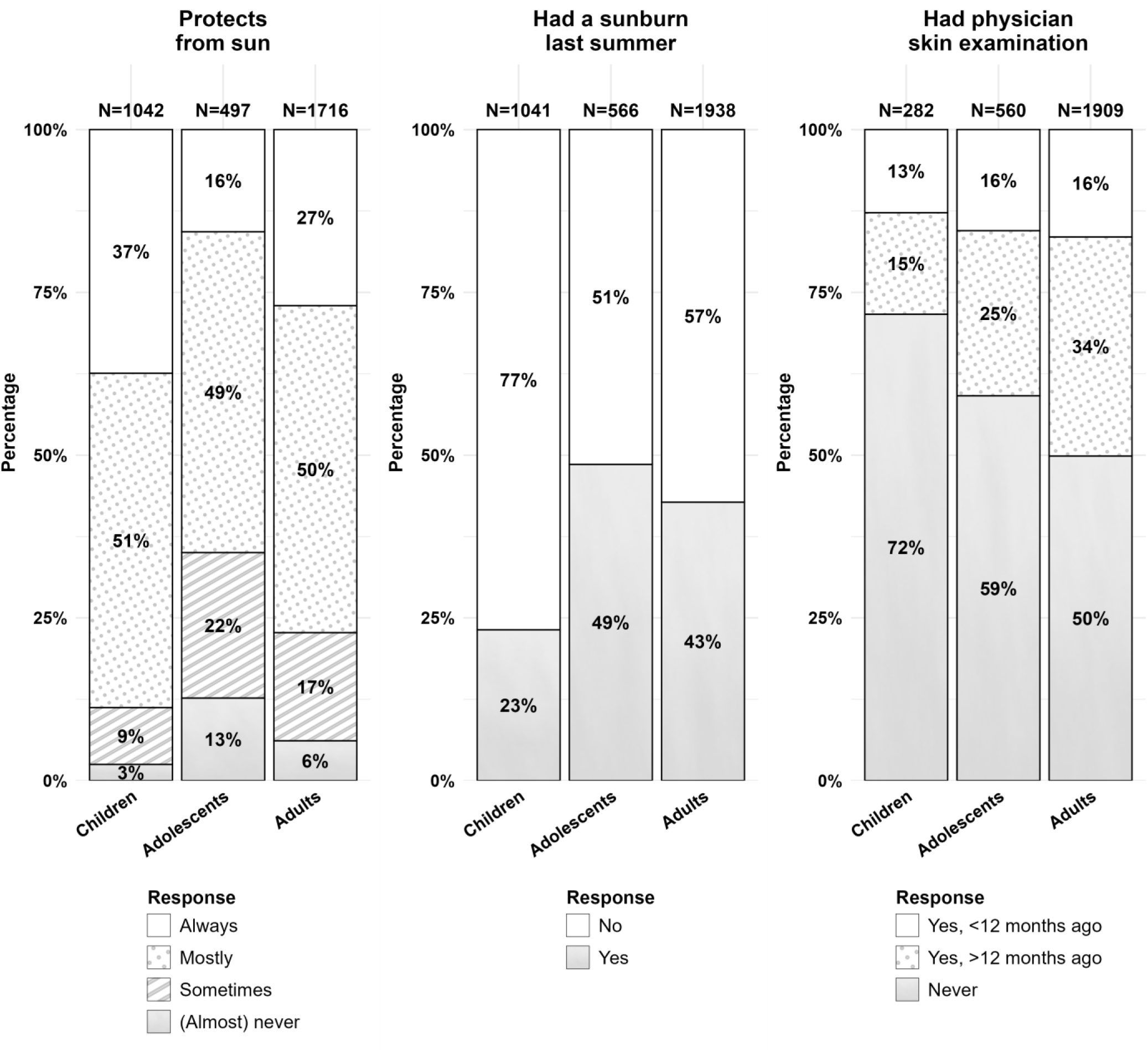
Prevalence of sun protection, sunburn, and physician skin examinations among childhood cancer survivors stratified by age group

**Figure 2.**
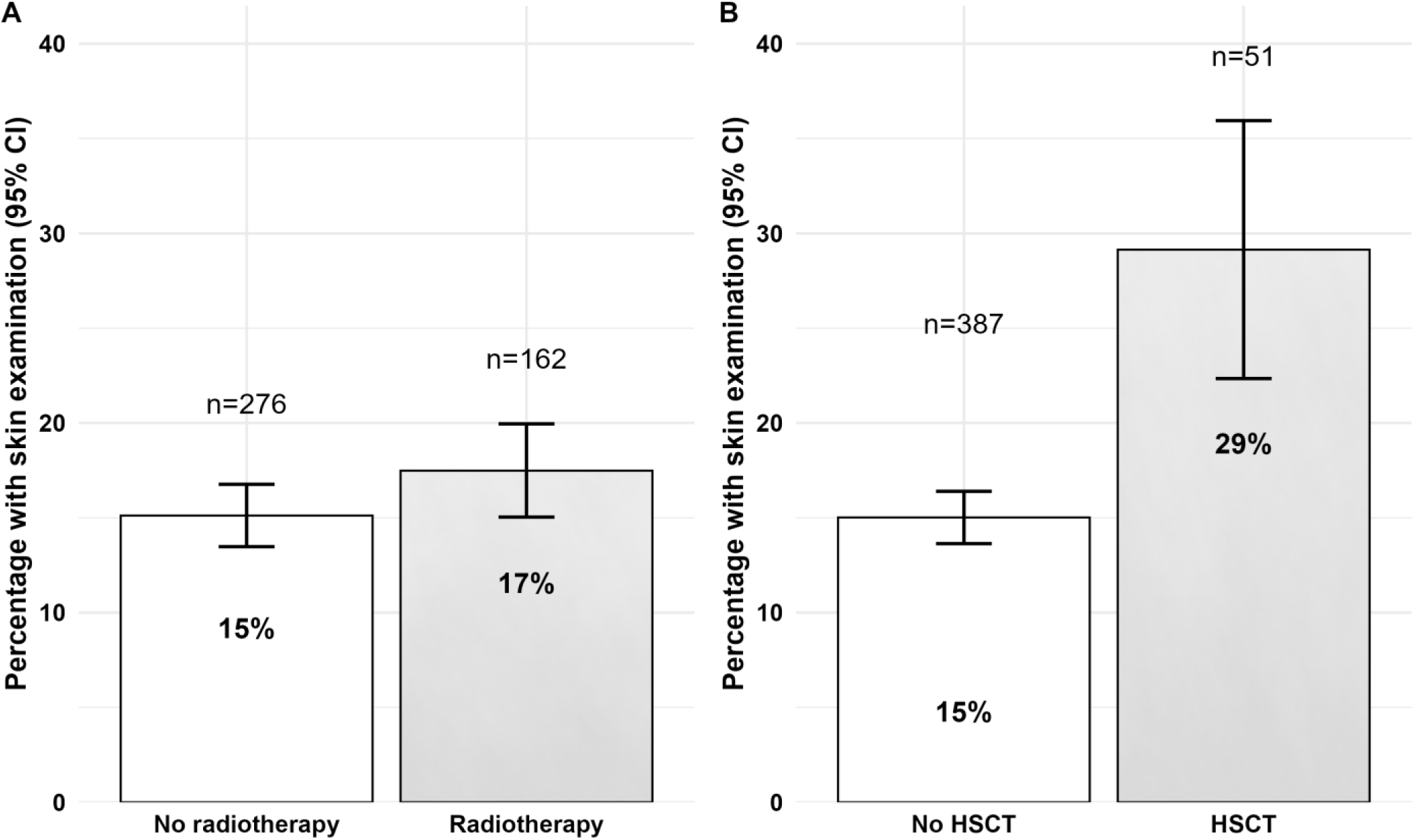
Compliance with physician skin examination attendance within the last 12 months as recommended by the Children’s Oncology Group for childhood cancer survivors treated with radiotherapy (Panel A) and HSCT (Panel B)

### Factors associated with sun protection and sunburn

*Table 2* shows results from logistic regression analyses, *Supplementary Table 5* results from Poisson regression. Across age groups, good sun protection was less likely among survivors with a migration background, whose parents had only primary education, and among survivors born more recently, indicating birth cohort effect. Independent of birth cohort, among children, higher age was associated with less sun protection (OR = 0.78, 95%CI 0.71-0.85). Male adolescent (OR = 0.37, 95%CI 0.24-0.56) and adult survivors (OR = 0.51, 95%CI 0.40-0.65) were less likely to protect from sun than females. Radiotherapy and HSCT treatment were unrelated to sun protection.

**Table 2.** Factors associated with sun protection, sunburn, and physician skin examinations in multivariable logistic regression in child, adolescents, and adults CCS.

|  | Children (5-15 years) |  |  | Adolescents (16-19 years) |  |  | Adults (≥20 years) |  |  |
| --- | --- | --- | --- | --- | --- | --- | --- | --- | --- |
|  | OR | 95%CI | p | OR | 95%CI | p | OR | 95%CI | p |
| **Sun protection** |  |  |  |  |  |  |  |  |  |
|  | N = 1,013 |  |  | N = 476 |  |  | N = 1,626 |  |  |
| Age at study | 0.73 | 0.66;0.82 | <0.001 | 0.91 | 0.77;1.08 | 0.294 | 1.01 | 0.98;1.04 | 0.543 |
| Male<sup>a</sup> | 0.72 | 0.47;1.09 | 0.122 | 0.38 | 0.25;0.57 | <0.001 | 0.51 | 0.40;0.65 | <0.001 |
| Migration background<sup>b</sup> | 0.64 | 0.39;1.07 | 0.083 | 0.60 | 0.35;1.01 | 0.053 | 0.58 | 0.43;0.79 | 0.001 |
| French/Italian language region<sup>c</sup> | 1.25 | 0.80;2.00 | 0.331 | 1.05 | 0.67;1.66 | 0.822 | 1.01 | 0.77;1.32 | 0.967 |
| Parental education<sup>d</sup> |  |  |  |  |  |  |  |  |  |
| Secondary | 1.98 | 0.98;3.93 | 0.053 | 1.34 | 0.58;3.11 | 0.489 | 1.30 | 0.87;1.94 | 0.193 |
| Tertiary | 1.98 | 0.99;3.84 | 0.048 | 1.92 | 0.79;4.65 | 0.149 | 1.55 | 1.01;2.36 | 0.042 |
| Radiotherapy<sup>e</sup> | 1.34 | 0.78;2.45 | 0.310 | 1.58 | 1.01;2.52 | 0.050 | 1.20 | 0.93;1.56 | 0.155 |
| HSCT<sup>f</sup> | 1.09 | 0.50;2.75 | 0.842 | 1.59 | 0.67;4.11 | 0.314 | 1.01 | 0.60;1.77 | 0.962 |
| Birth year | 0.94 | 0.89;0.99 | 0.027 | 0.95 | 0.91;1.00 | 0.047 | 0.97 | 0.94;0.99 | 0.004 |
| **Sun burn** |  |  |  |  |  |  |  |  |  |
|  | N = 1,013 |  |  | N = 542 |  |  | N = 1,837 |  |  |
| Age at study | 1.28 | 1.18;1.38 | <0.001 | 1.03 | 0.89;1.18 | 0.719 | 1.01 | 0.99;1.03 | 0.411 |
| Male<sup>a</sup> | 0.98 | 0.73;1.33 | 0.916 | 0.73 | 0.52;1.03 | 0.076 | 1.05 | 0.86;1.27 | 0.632 |
| Migration background<sup>b</sup> | 0.53 | 0.34;0.81 | 0.004 | 0.72 | 0.44;1.17 | 0.187 | 0.48 | 0.36;0.63 | <0.001 |
| French/Italian language region<sup>c</sup> | 0.95 | 0.68;1.31 | 0.740 | 1.11 | 0.75;1.63 | 0.605 | 0.85 | 0.68;1.05 | 0.137 |
| Parental education<sup>d</sup> |  |  |  |  |  |  |  |  |  |
| Secondary | 1.09 | 0.58;2.13 | 0.801 | 1.08 | 0.48;2.46 | 0.853 | 1.18 | 0.82;1.70 | 0.379 |
| Tertiary | 0.90 | 0.49;1.76 | 0.759 | 1.33 | 0.58;3.12 | 0.501 | 1.47 | 1.01;2.14 | 0.044 |
| Radiotherapy<sup>e</sup> | 0.67 | 0.43;1.01 | 0.065 | 0.63 | 0.43;0.93 | 0.022 | 0.77 | 0.63;0.94 | 0.012 |
| HSCT<sup>f</sup> | 0.61 | 0.30;1.14 | 0.143 | 1.45 | 0.72;2.95 | 0.303 | 0.62 | 0.39;0.95 | 0.031 |
| Birth year | 1.14 | 1.10;1.19 | <0.001 | 1.04 | 0.99;1.08 | 0.101 | 1.07 | 1.05;1.09 | <0.001 |
| **Physician skin examination** |  |  |  |  |  |  |  |  |  |
|  | N = 275 |  |  | N = 538 |  |  | N = 1,807 |  |  |
| Age at study | 0.27 | 0.07;0.94 | 0.044 | 1.02 | 0.83;1.24 | 0.874 | 1.04 | 1.01;1.07 | 0.019 |
| Male<sup>a</sup> | 0.97 | 0.45;2.10 | 0.942 | 1.38 | 0.85;2.27 | 0.194 | 0.65 | 0.50;0.83 | 0.001 |
| Migration background<sup>b</sup> | 0.87 | 0.33;2.08 | 0.757 | 1.28 | 0.67;2.36 | 0.435 | 1.35 | 0.95;1.89 | 0.086 |
| French/Italian language region<sup>c</sup> | 2.21 | 0.99;4.93 | 0.051 | 1.08 | 0.63;1.81 | 0.773 | 1.39 | 1.06;1.82 | 0.016 |
| Parental education<sup>g</sup> |  |  |  |  |  |  |  |  |  |
| Secondary |  |  |  | 1.55 | 0.51;5.80 | 0.471 | 0.96 | 0.62;1.50 | 0.844 |
| Tertiary | 3.27 | 1.20;10.79 | 0.032 | 1.58 | 0.51;6.09 | 0.460 | 1.28 | 0.82;2.02 | 0.289 |
| Radiotherapy<sup>e</sup> | 2.62 | 0.94;6.86 | 0.054 | 1.15 | 0.67;1.93 | 0.596 | 1.01 | 0.77;1.32 | 0.935 |
| HSCT<sup>f</sup> | 3.75 | 1.39;9.83 | 0.008 | 2.06 | 0.87;4.55 | 0.083 | 2.07 | 1.29;3.27 | 0.002 |
| Birth year | 0.31 | 0.08;1.09 | 0.072 | 1.00 | 0.94;1.06 | 0.927 | 1.01 | 0.99;1.04 | 0.339 |

<sup>a</sup> reference: female, <sup>b</sup> reference: no migration background, <sup>c</sup> reference: German language region, <sup>d</sup> reference: primary education, <sup>e</sup> reference: no radiotherapy; <sup>f</sup> reference: no HSCT, <sup>g</sup> due to the lower n in this analysis, we combined primary and secondary education into in category for children; for adolescents and adults, we kept it separate; HSCT = hematopoietic stem cell transplantation

Similar to sun protection, more recent birth cohorts were more likely to report sunburn. Independent of birth cohort, among children, higher age was associated with higher sunburn odds (OR = 1.28, 95%CI 1.18-1.38). Survivors with migration background were less likely to report sunburn across children and adults. Sunburn was less likely among pediatric (OR = 0.67, 95%CI 0.43-1.01), adolescent (OR = 0.63, 95%CI 0.43-0.93), and adult (OR = 0.77, 95%CI 0.63-0.94) survivors treated with radiotherapy, and among adult HSCT recipients (OR = 0.62, 95%CI 0.39-0.95).

### Factors associated with PSE

HSCT pediatric (OR = 3.75, 95%CI 1.39-9.83), adolescent (OR = 2.06, 95%CI = 0.87-4.55), and adult (OR = 2.07, 95%CI 1.29-3.27) recipients were more likely to have had a PSE within the last 12 months. For radiotherapy, this only applied to pediatric survivors (OR = 2.62, 95% CI 0.94–6.86) (*Table 2*). Pediatric (OR = 2.21, 95%CI 0.99-4.95) and adult (OR=1.39, 95%CI 1.06-1.82) survivors in the French or Italian language region were more likely to have had a PSE, as were pediatric survivors whose parents had tertiary education. PSE was unrelated to birth year.

### Sensitivity analyses

Across all outcomes, sensitivity analyses yielded similar results. When including skin type, the direction of associations remained unchanged: darker skin type was linked to lower likelihood of sun protection and sunburn but was unrelated to PSE. Results were also stable when replacing birth year with year of study participation (*Supplementary Tables 6-8*).

## Discussion

Despite their elevated skin cancer risk, one in five childhood cancer survivors in our study reported insufficient sun protection, and one in three experienced a sunburn during the past year. Sun protection was poorer and sunburn more frequent among younger generations.

Survivors who received radiotherapy or HSCT did not protect substantially better from the sun than those without these treatments. Only 18% of survivors treated with radiotherapy and 29% of those treated with HSCT reported a physician skin examination (PSE) within the past year, as recommended in the COG guidelines.

Compared with previous CCS studies, sun protection was relatively high in our study. In the North-American Childhood Cancer Survivor Study with predominantly adult survivors, 67% reported regular sunscreen use and 41% report wearing protective clothing.^13^ In smaller survivor studies, only 23%-59% apply sun safety measures such as using sunscreen, wearing protective clothing, or seeking shade.^15,20,30^ Differences may partly reflect measurement approaches: we assessed sun protection as a composite of behaviors (sunscreen use, shading, protective clothing), whereas other studies evaluated single behaviors.^13,15,30^ In the general Swiss population, 54-80% of children^31–33^ and 77% of young adults use sunscreen,^34^ which is slightly lower or similar to our study. Sunburn was less common among pediatric survivors than among children in the general population (40–60%),^31,32^ which may reflect better protection through parents after their child’s cancer. Sunburn prevalence among adult survivors was comparable to adults in Germany (49%),^35^ which may reflect decreased awareness compared to children due to longer time since treatment and less presence of their cancer history in everyday life (e.g., less follow-up examinations).^36^ Sun protection was lowest and sunburn prevalence highest among adolescent survivors. This finding is consistent with previous evidence highlighting adolescence as a vulnerable stage for poor health behaviors.^37^

Across age groups, survivors born more recently were less likely to use sun protection and more likely to report sunburn. This generational pattern may reflect reduced public awareness of skin cancer prevention compared to the 1990s, when there was considerable media attention on ozone layer depletion and UV-related risks.^38^ Younger generations are additionally exposed to misinformation about sunscreen safety, including vitamin D concerns^39^ and alleged carcinogenic side effects,^40^ combined with appearance norms favoring tanning, amplified through social media.^41^ This trend requires close monitoring. In our study, female adolescent and adult survivors were more likely to protect from sun. This is consistent with previous findings,^13,16,20^ mirroring general evidence that women tend to be more health-conscious and seek preventive information more actively.^42^ Survivors with migration background were less likely to protect from sun and less likely to report sunburn. This may be partially explained by skin phototype (30% of those with migration background report a skin type 4-6 compared to 18% of those without migration background), but also by culturally different sunseeking and sun protection habits.^18^ Radiotherapy was not associated with better sun protection, aligning with previous findings,^15^ yet was linked to lower sunburn odds in adolescents and adults, as was HSCT among adults. This points to sun avoidance rather than sun protection.

Only 16% of survivors reported a PSE within the past 12 months, which is slightly higher than the 13% observed in the general Swiss population.^43^ Compliance with the COG’s latest PSE recommendations was higher among HSCT recipients (29%) than among those treated with radiotherapy (17%). The latter figure is consistent with 11-31% reported in North American cohorts.^44–46^ Low PSE attendance may result from limited knowledge of prior treatments and associated risks.^44,47^ While still low, higher attendance among HSCT survivors likely reflects their ongoing, multidisciplinary follow-up care due to greater morbidity burden^48,49^ and frequent dermatological problems,^50^ which may increase screening opportunities.Further investigations, such as in interviewing survivors about barriers they encounter to attend skin examinations, can provide valuable insights. Older adults were more likely to report PSE attendance, potentially reflecting increasing health awareness with older age.^51^ Regional differences were also evident: survivors from French- and Italian-speaking regions reported higher PSE attendance. This may relate to differences in preventive care culture since those regions have longer-standing organized screening programs such as mammography.^52^

This study provides comprehensive, population-based estimates of sun protection, sunburn, and PSE among CCS across the full age spectrum and over two decades of data collection. It extends prior work by assessing adherence to COG recommendations after both radiotherapy and HSCT. Limitations include self-reporting, which may introduce a social desirability bias. Incomplete availability of some outcomes and exposures, particularly PSE in children and skin type, limits the study’s statistical power. We did not assess actual sun exposure and skin self-examination, which the COG recommends monthly and which can reduce melanoma mortality by 63%.^53^ Our cohort is predominantly of Western European ancestry, which may limit generalizability to more diverse populations.

Our findings highlight the need for system-level strategies to embed sun protection and skin cancer screening into survivorship care. Integrating dermatology referrals and annual reminders for radiation or HSCT recipients within survivorship care plans^54^ (e.g., Survivorship Passport) as well as patient or provider activation regarding skin cancer risk after childhood cancer^55^ can improve knowledge and recommenadtion adherence. Education efforts should emphasize the importance of sun protection for all survivors.^55^

Healthcare practitioners engaged in follow-up care should support survivors to attend yearly PSE, particularly after radiotherapy or HSCT, and encourage consistent sun protection, especially among adolescents and male survivors, and younger generations.

## Data Availability

We accessed the data that support the information of this manuscript via secured servers of the Institute of Social and Preventive Medicine at the University of Bern. Individual-level, fully anonymized, sensitive data can only be made available for researchers who fulfil the legal requirements. Requests for data from the Childhood Cancer Registry of Switzerland must be directed to this organization (https://www.childhoodcancerregistry.ch). Requests for data from the Swiss Childhood Cancer Survivor Study (SCCSS) should be communicated to the study lead, Claudia E. Kuehni.

## Précis

Sun protection is poor among childhood cancer survivors, particularly among younger generations. Only one in five treated with radiotherapy or HSCT meets current recommendations for skin cancer screening. This highlights the need to systematically integrate sun safety education and skin cancer screening into follow-up care.

## Acknowledgement

We thank all survivors for participating in our study, the study team of the Childhood Cancer Research Group, the data managers of the Swiss Paediatric Oncology Group, and the team of the Swiss Childhood Cancer Registry.

## Ethics approval statement

Ethical approval was granted by the ethics committee of the canton of Bern, Switzerland (KEK-BE: 166/2014 and 2021-01462).

## Clinical trial registration

Clinicaltrials.gov (NCT03297034)

---

**Supplementary Figure 1.**
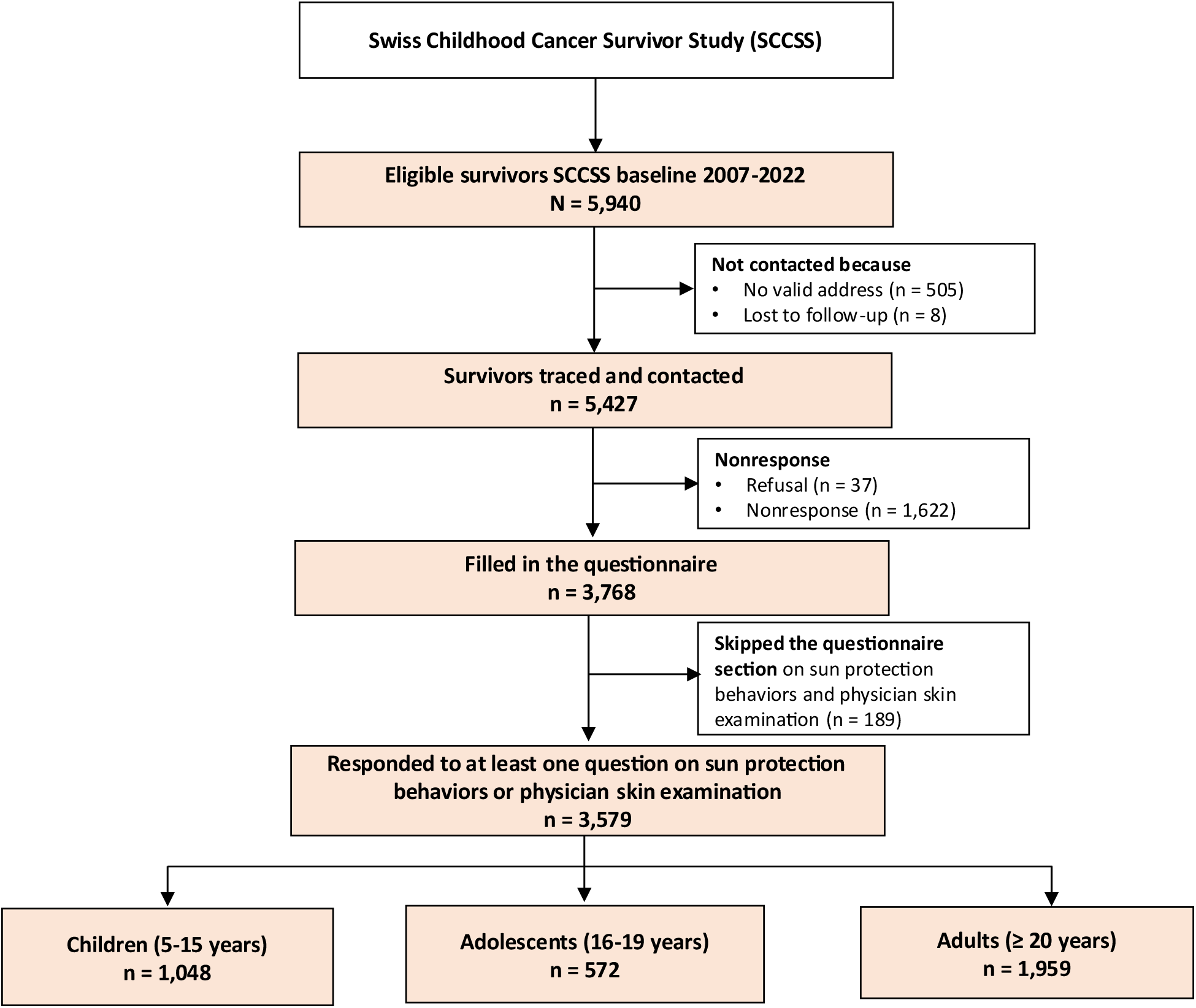
Study population tree of the SCCSS study on sun protection behaviors and physician skin examination

**Supplementary Table 1.** Overview sun protection behaviors and physician skin examination questions, response options, and categorization.

| Outcome variables | Question | Response option | Coding for regression analysis |
| --- | --- | --- | --- |
| Sun protection | How consequentially to you protect yourself (for parent questionnaire: your child) from sun? For example, applying sunscreen, wearing clothing and/or a hat, shading, considering day time for one's sun exposure | 1: Always<br>2: Most of the time<br>3: Every now and then<br>4: Never | 1 and 2: Yes<br>3 and 4: No |
| Sunburn | Have you (for parent questionnaire: your child) had one or more sunburns the previous summer? | 1: No<br>2: Yes | 1: No<br>2: Yes |
| Preventive skin examinations | Have you (your child) ever had your skin or moles medically examined? | 1: Yes, less than 12 months ago<br>2: Yes, more than 12 months ago<br>3: No, never | 1 and 2: Yes<br>3: No |

*Please note: Not all questions were included in all surveys or in all age groups. <sup>a</sup> In earlier surveys, response options were only "yes" and "no"*

**Supplementary Table 2.** Model fit indices of multivariable logistic regression models including birth year compared to multivariable logistic regression models including year of study

| Model | AIC | BIC |
| --- | --- | --- |
| **Children (5-15 years)** |  |  |
| Sun protection with birth year | 658 | 708 |
| Sun protection with year of study | 659 | 713 |
| Sunburn with birth year | 1,055 | 1,104 |
| Sunburn with year of study | 1,057 | 1,111 |
| Skin exam with birth year | 200 | 232 |
| Skin exam with study year | 201 | 230 |
| **Adolescents (16-19 years)** |  |  |
| Sun protection with birth year | 593 | 635 |
| Sun protection with year of study | 584 | 630 |
| Sunburn with birth year | 753 | 796 |
| Sunburn with year of study | 756 | 808 |
| Skin exam with birth year | 466 | 509 |
| Skin exam with year of study | 470 | 521 |
| **Adults ($\geq 20$ years)** |  |  |
| Sun protection with birth year | 1,675 | 1,729 |
| Sun protection with year of study | 1,676 | 1,736 |
| Sunburn with birth year | 2,371 | 2,426 |
| Sunburn with year of study | 2,375 | 2,441 |
| Skin exam with birth year | 1,577 | 1,632 |
| Skin exam with year of study | 1,579 | 1,645 |

**Supplementary Table 3.** Prevalence of sun protection behaviors and physician skin examinations among childhood cancer survivors.

|  | Children (5-15 years) |  | Adolescents (16-19 years) |  | Adults (≥ 20 years) |  | Overall |  |
| --- | --- | --- | --- | --- | --- | --- | --- | --- |
|  | N | 95%CI | N | 95%CI | N | 95%CI | N | 95%CI |
| *Sun protection* | **n = 1,048** |  | **n = 499** |  | **n = 1,727** |  | **n = 3,274** |  |
| Always | 390 (37%) | 34-40 | 78 (16%) | 13-19 | 464 (27%) | 25-29 | 932 (28%) | 27-30 |
| Mostly | 535 (51%) | 48-54 | 245 (49%) | 45-53 | 862 (50%) | 48-52 | 1642 (50%) | 48-52 |
| Sometimes | 91 (9%) | 7-11 | 111 (22%) | 19-26 | 285 (17%) | 15-18 | 487 (15%) | 14-16 |
| (Almost) never | 26 (3%) | 2-4 | 63 (13%) | 10-16 | 105 (6%) | 5-7 | 194 (6%) | 5-7 |
| Missing | 6 (1%) |  | 2 (<1%) |  | 11 (1%) |  | 19 (<1%) |  |
| *Sun protection (dichotomized)* | **n = 1,048** |  | **n = 499** |  | **n = 1,727** |  | **n = 3,274** |  |
| No | 117 (11%) | 9-13 | 174 (35%) | 31-39 | 390 (23%) | 21-25 | 681 (21%) | 19-22 |
| Yes | 925 (89%) | 86-90 | 323 (65%) | 60-69 | 1326 (77%) | 75-79 | 2574 (79%) | 77-80 |
| Missing | 6 (1%) |  | 2 (0.4%) |  | 11 (1%) |  | 19 (<1%) |  |
| *Sunburn(s) last summer* | **n = 1,048** |  | **n = 572** |  | **n = 1,959** |  | **n = 3,579** |  |
| No | 800 (77%) | 74-79 | 291 (51%) | 47-55 | 1109 (57%) | 54-59 | 2200 (62%) | 60-63 |
| Yes | 241 (23%) | 21-26 | 275 (49%) | 44-52 | 829 (43%) | 40-45 | 1345 (38%) | 36-39 |
| Missing | 7 (1%) |  | 6 (1%) |  | 21 (1%) |  | 34 (1%) |  |
| *Physician skin examination* | **n = 284** |  | **n = 572** |  | **n = 1,959** |  | **n = 2,815** |  |
| Yes, < 12 months ago | 36 (13%) | 9-17 | 87 (16%) | 12-18 | 315 (16%) | 15-18 | 438 (16%) | 14-17 |
| Yes, ≥ 12 months ago | 44 (16%) | 12-20 | 142 (25%) | 21-29 | 642 (33%) | 31-25 | 828 (30%) | 28-31 |
| No, never | 202 (72%) | 66-76 | 331 (59%) | 54-62 | 952 (50%) | 46-51 | 1485 (54%) | 51-55 |
| Missing | 2 (1%) |  | 12 (2%) |  | 50 (3%) |  | 64 (2%) |  |
| *Physician skin examination within the last 12 months* | **n = 284** |  | **n = 572** |  | **n = 1,959** |  | **n = 2,815** |  |
| No | 246 (87%) | 82-90 | 473 (84%) | 79-86 | 1594 (84%) | 80-83 | 2313 (82%) | 81-84 |
| Yes | 36 (13%) | 9-17 | 87 (16%) | 12-18 | 315 (16%) | 15-18 | 438 (16%) | 14-17 |
| Missing | 2 (1%) |  | 12 (2%) |  | 50 (3%) |  | 64 (2%) |  |

Please note: Not all questions were asked in all surveys or in all age categories so that we report the available *n* for each variable.

**Supplementary Table 4.**
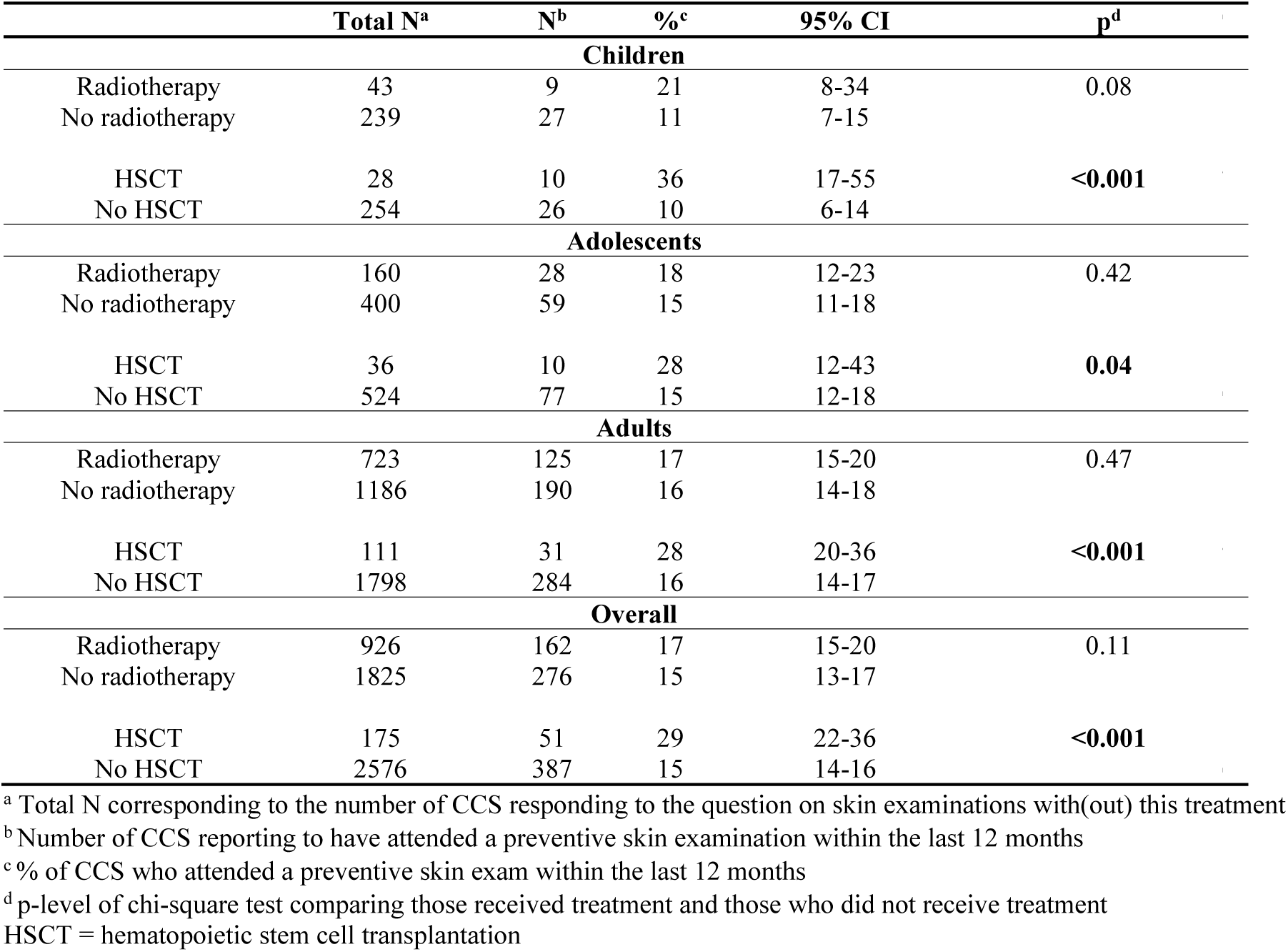
Proportion of childhood cancer survivors who attended a physician skin examination within the last 12 months as recommended by the Children’s Oncology Group 2023 Follow-Up Care Guidelines stratified by radiotherapy and HSCT exposure

**Supplementary Table 5.** Factors associated with sun protection, sunburn, and physician skin examination in modified Poisson regression in child, adolescents, and adults childhood cancer survivors

|  | Children (5-15 years) |  |  | Adolescents (16-19 years) |  |  | Adults (≥20 years) |  |  |
| --- | --- | --- | --- | --- | --- | --- | --- | --- | --- |
|  | RR | 95%CI | p | RR | 95%CI | p | RR | 95%CI | p |
| **Sun protection** |  |  |  |  |  |  |  |  |  |
|  | **N = 1,013** |  |  | **N = 476** |  |  | **N = 1,626** |  |  |
| Age at study | 0.97 | 0.96;0.98 | <0.001 | 0.97 | 0.92;1.03 | 0.301 | 1.00 | 0.99;1.01 | 0.967 |
| Male<sup>a</sup> | 0.97 | 0.93;1.01 | 0.143 | 0.73 | 0.64;0.84 | <0.001 | 0.87 | 0.82;0.91 | <0.001 |
| Migration background<sup>b</sup> | 0.95 | 0.89;1.01 | 0.080 | 0.83 | 0.66;1.03 | 0.093 | 0.87 | 0.79;0.95 | 0.001 |
| French/Italian language region<sup>c</sup> | 1.02 | 0.98;1.07 | 0.350 | 1.02 | 0.88;1.18 | 0.783 | 1.00 | 0.94;1.06 | 0.967 |
| Parental education<sup>d</sup> |  |  |  |  |  |  |  |  |  |
| Secondary | 1.12 | 1.00;1.26 | 0.059 | 1.13 | 0.78;1.64 | 0.522 | 1.06 | 0.96;1.18 | 0.223 |
| Tertiary | 1.12 | 1.00;1.26 | 0.058 | 1.27 | 0.86;1.85 | 0.226 | 1.10 | 0.99;1.22 | 0.065 |
| Radiotherapy<sup>e</sup> | 1.03 | 0.98;1.09 | 0.287 | 1.15 | 1.01;1.31 | 0.037 | 1.04 | 0.99;1.10 | 0.146 |
| HSCT<sup>f</sup> | 1.01 | 0.93;1.08 | 0.874 | 1.14 | 0.92;1.42 | 0.226 | 1.00 | 0.90;1.12 | 0.961 |
| Birth year | 0.99 | 0.99;1.00 | 0.048 | 0.98 | 0.96;1.00 | 0.068 | 0.99 | 0.98;1.00 | 0.007 |
| **Sunburn** |  |  |  |  |  |  |  |  |  |
|  | **N = 1,013** |  |  | **N = 542** |  |  | **N = 1,837** |  |  |
| Age at study | 1.20 | 1.13;1.26 | <0.001 | 1.01 | 0.94;1.09 | 0.730 | 1.00 | 0.99;1.01 | 0.971 |
| Male<sup>a</sup> | 0.99 | 0.80;1.22 | 0.914 | 0.85 | 0.72;1.02 | 0.075 | 1.02 | 0.93;1.13 | 0.636 |
| Migration background<sup>b</sup> | 0.63 | 0.46;0.87 | 0.005 | 0.84 | 0.65;1.10 | 0.212 | 0.65 | 0.54;0.78 | <0.001 |
| French/Italian language region<sup>c</sup> | 0.96 | 0.76;1.22 | 0.754 | 1.05 | 0.87;1.27 | 0.603 | 0.92 | 0.82;1.03 | 0.164 |
| Parental education<sup>d</sup> |  |  |  |  |  |  |  |  |  |
| Secondary | 1.06 | 0.66;1.71 | 0.802 | 1.05 | 0.66;1.68 | 0.837 | 1.12 | 0.89;1.41 | 0.343 |
| Tertiary | 0.93 | 0.58;1.49 | 0.766 | 1.16 | 0.72;1.88 | 0.532 | 1.25 | 0.99;1.59 | 0.058 |
| Radiotherapy<sup>e</sup> | 0.74 | 0.54;1.02 | 0.068 | 0.79 | 0.63;0.98 | 0.029 | 0.86 | 0.77;0.96 | 0.010 |
| HSCT<sup>f</sup> | 0.69 | 0.41;1.17 | 0.165 | 1.19 | 0.88;1.60 | 0.259 | 0.76 | 0.58;1.00 | 0.047 |
| Birth year | 1.10 | 1.07;1.14 | <0.001 | 1.02 | 1.00;1.04 | 0.088 | 1.03 | 1.02;1.04 | <0.001 |
| **Physician skin examination** |  |  |  |  |  |  |  |  |  |
|  | **N = 275** |  |  | **N = 538** |  |  | **N = 1,807** |  |  |
| Age at study | 0.34 | 0.11;1.06 | 0.063 | 1.01 | 0.87;1.19 | 0.861 | 1.03 | 1.01;1.06 | 0.018 |
| Male<sup>a</sup> | 0.96 | 0.54;1.73 | 0.899 | 1.32 | 0.87;1.99 | 0.190 | 0.70 | 0.57;0.86 | 0.001 |
| Migration background<sup>b</sup> | 0.92 | 0.41;2.06 | 0.835 | 1.23 | 0.74;2.04 | 0.421 | 1.27 | 0.97;1.67 | 0.081 |
| French/Italian language region<sup>c</sup> | 1.85 | 1.01;3.42 | 0.048 | 1.07 | 0.69;1.64 | 0.762 | 1.31 | 1.05;1.63 | 0.017 |
| Parental education<sup>g</sup> |  |  |  |  |  |  |  |  |  |
| Secondary |  |  |  | 1.45 | 0.54;3.85 | 0.461 | 0.96 | 0.68;1.37 | 0.839 |
| Tertiary | 2.70 | 1.03;7.11 | 0.044 | 1.47 | 0.54;4.04 | 0.451 | 1.22 | 0.85;1.74 | 0.281 |
| Radiotherapy<sup>e</sup> | 2.16 | 1.10;4.26 | 0.026 | 1.13 | 0.72;1.76 | 0.601 | 1.01 | 0.81;1.25 | 0.935 |
| HSCT<sup>f</sup> | 2.61 | 1.39;4.89 | 0.003 | 1.79 | 0.95;3.34 | 0.070 | 1.75 | 1.24;2.46 | 0.002 |
| Birth year | 0.39 | 0.12;1.21 | 0.103 | 1.00 | 0.95;1.06 | 0.923 | 1.01 | 0.99;1.03 | 0.331 |

<sup>a</sup> reference: female, <sup>b</sup> reference: no migration background, <sup>c</sup> reference: German language region, <sup>d</sup> reference: primary education, <sup>e</sup> reference: no radiotherapy; <sup>f</sup> reference: no HSCT, <sup>g</sup> due to the lower n in this analysis, we combined primary and secondary education into in category for children; for adolescents and adults, we kept it separate, HSCT = hematopoietic stem cell transplantation

**Supplementary Table 6.** Skin type descriptives of pediatric, adolescent, and adult childhood cancer survivors.

|  | Children (5-15<br>years)<br>n = 598 | Adolescents<br>(16-19 years)<br>n = 572 | Adults (≥ 20 years)<br>n = 470 | Overall<br>N = 1,640 |
| --- | --- | --- | --- | --- |
| *Skin type 5 levels* |  |  |  |  |
| Skin type 1 | 25 (4%) | 18 (3%) | 20 (4%) | 63 (4%) |
| Skin type 2 | 159 (27%) | 142 (25%) | 125 (27%) | 426 (26%) |
| Skin type 3 | 282 (48%) | 285 (51%) | 237 (51%) | 804 (49%) |
| Skin type 4 | 112 (19%) | 107 (19%) | 80 (17%) | 299 (18%) |
| Skin type 5 | 14 (2%) | 12 (2%) | 5 (1%) | 31 (2%) |
| Missing | 6 (1%) | 8 (1%) | 3 (1%) | 17 (1%) |
| *Skin type 3 levels (for*<br>*analysis)* |  |  |  |  |
| Skin type 1-2 | 184 (31%) | 160 (28%) | 145 (31%) | 489 (30%) |
| Skin type 3 | 282 (48%) | 285 (51%) | 237 (51%) | 804 (50%) |
| Skin type 4-5 | 126 (21%) | 119 (21%) | 85 (18%) | 330 (20%) |
| Missing | 6 (1%) | 8 (1%) | 3 (1%) | 17 (1%) |

**Supplementary Table 7.** Sensitivity analysis: Factors associated with sun protection, sunburn, and physician skin examinations in multivariable logistic regression in child, adolescents, and adults childhood cancer survivors, including skin type

|  | Children (5-15 years) |  |  | Adolescents (16-19 years) |  |  | Adults (≥20 years) |  |  |
| --- | --- | --- | --- | --- | --- | --- | --- | --- | --- |
|  | OR | 95%CI | p | OR | 95%CI | p | OR | 95%CI | p |
| **Sun protection** |  |  |  |  |  |  |  |  |  |
|  | **N = 576** |  |  | **N = 470** |  |  | **N = 224** |  |  |
| Age at study | 0.80 | 0.71;0.89 | <0.001 | 0.95 | 0.80;1.13 | 0.538 | 1.03 | 0.98;1.09 | 0.224 |
| Male<sup>a</sup> | 0.97 | 0.56;1.67 | 0.908 | 0.42 | 0.27;0.64 | <0.001 | 0.66 | 0.34;1.25 | 0.203 |
| Migration background<sup>b</sup> | 0.80 | 0.42;1.55 | 0.492 | 0.66 | 0.38;1.15 | 0.142 | 0.37 | 0.15;0.93 | 0.035 |
| French/Italian language region<sup>c</sup> | 0.84 | 0.46;1.58 | 0.573 | 0.87 | 0.55;1.40 | 0.572 | 1.56 | 0.80;3.16 | 0.202 |
| Parental education<sup>d</sup> |  |  |  |  |  |  |  |  |  |
| Secondary | 2.71 | 0.98;7.30 | 0.050 | 1.01 | 0.43;2.39 | 0.973 | 0.93 | 0.26;3.13 | 0.912 |
| Tertiary | 1.89 | 0.73;4.65 | 0.176 | 1.31 | 0.53;3.21 | 0.559 | 1.29 | 0.37;4.24 | 0.682 |
| Radiotherapy<sup>e</sup> | 2.14 | 0.96;5.47 | 0.083 | 1.71 | 1.08;2.74 | 0.025 | 1.55 | 0.78;3.18 | 0.215 |
| HSCT<sup>f</sup> | 0.84 | 0.32;2.64 | 0.746 | 1.53 | 0.64;4.03 | 0.361 | 0.71 | 0.21;2.50 | 0.578 |
| Skin type (ref. 1-2) |  |  |  |  |  |  |  |  |  |
| Skin type 3 | 0.25 | 0.10;0.57 | 0.002 | 0.48 | 0.28;0.81 | 0.008 | 0.59 | 0.28;1.23 | 0.166 |
| Skin type 4-6 | 0.12 | 0.04;0.28 | <0.001 | 0.24 | 0.12;0.44 | <0.001 | 0.21 | 0.09;0.51 | 0.001 |
| **Sunburn** |  |  |  |  |  |  |  |  |  |
|  | **N = 576** |  |  | **N = 537** |  |  | **N = 445** |  |  |
| Age at study | 1.14 | 1.06;1.23 | 0.001 | 1.02 | 0.88;1.18 | 0.806 | 0.93 | 0.91;0.96 | <0.001 |
| Male<sup>a</sup> | 1.03 | 0.70;1.52 | 0.866 | 0.82 | 0.57;1.18 | 0.286 | 1.05 | 0.70;1.57 | 0.819 |
| Migration background<sup>b</sup> | 0.60 | 0.36;0.99 | 0.050 | 0.93 | 0.56;1.55 | 0.783 | 0.40 | 0.22;0.72 | 0.002 |
| French/Italian language region<sup>c</sup> | 0.71 | 0.46;1.09 | 0.118 | 0.87 | 0.58;1.30 | 0.503 | 1.19 | 0.77;1.86 | 0.433 |
| Parental education<sup>d</sup> |  |  |  |  |  |  |  |  |  |
| Secondary | 1.00 | 0.45;2.35 | 0.992 | 0.81 | 0.34;1.93 | 0.628 | 0.82 | 0.37;1.82 | 0.622 |
| Tertiary | 0.67 | 0.30;1.53 | 0.330 | 1.02 | 0.42;2.47 | 0.973 | 1.18 | 0.53;2.62 | 0.685 |
| Radiotherapy<sup>e</sup> | 0.50 | 0.28;0.88 | 0.019 | 0.57 | 0.38;0.85 | 0.006 | 1.03 | 0.67;1.59 | 0.879 |
| HSCT<sup>f</sup> | 0.46 | 0.18;1.02 | 0.073 | 1.61 | 0.78;3.42 | 0.203 | 0.77 | 0.35;1.67 | 0.503 |
| Skin type (ref. 1-2) |  |  |  |  |  |  |  |  |  |
| Skin type 3 | 0.57 | 0.37;0.87 | 0.009 | 0.60 | 0.39;0.91 | 0.017 | 0.99 | 0.62;1.57 | 0.959 |
| Skin type 4-6 | 0.19 | 0.10;0.36 | <0.001 | 0.16 | 0.09;0.28 | <0.001 | 0.47 | 0.26;0.84 | 0.012 |
| **Physician skin examination** |  |  |  |  |  |  |  |  |  |
|  | **N = 270** |  |  | **N = 531** |  |  | **N = 442** |  |  |
| Age at study | 0.85 | 0.72;0.99 | 0.044 | 1.01 | 0.83;1.23 | 0.935 | 1.02 | 0.98;1.05 | 0.307 |
| Male<sup>a</sup> | 1.00 | 0.46;2.18 | 0.995 | 1.39 | 0.85;2.30 | 0.190 | 0.49 | 0.29;0.83 | 0.009 |
| Migration background<sup>b</sup> | 0.94 | 0.36;2.26 | 0.890 | 1.44 | 0.74;2.68 | 0.270 | 1.07 | 0.49;2.22 | 0.859 |
| French/Italian language region<sup>c</sup> | 2.43 | 1.06;5.61 | 0.036 | 0.99 | 0.57;1.69 | 0.983 | 1.18 | 0.67;2.05 | 0.557 |
| Parental education<sup>g</sup> |  |  |  |  |  |  |  |  |  |
| Secondary |  |  |  | 1.49 | 0.49;5.64 | 0.510 | 0.74 | 0.29;2.06 | 0.545 |
| Tertiary | 4.10 | 1.47;13.92 | 0.012 | 1.47 | 0.47;5.67 | 0.536 | 0.89 | 0.35;2.48 | 0.821 |
| Radiotherapy<sup>e</sup> | 2.55 | 0.91;6.78 | 0.064 | 1.12 | 0.66;1.88 | 0.663 | 1.00 | 0.57;1.74 | 0.993 |
| HSCT<sup>f</sup> | 3.63 | 1.33;9.56 | 0.010 | 2.02 | 0.85;4.44 | 0.091 | 1.88 | 0.72;4.50 | 0.170 |
| Skin type (ref. 1-2) |  |  |  |  |  |  |  |  |  |
| Skin type 3 | 0.57 | 0.23;1.41 | 0.219 | 0.59 | 0.34;1.04 | 0.066 | 0.95 | 0.54;1.70 | 0.866 |
| Skin type 4-6 | 1.40 | 0.49;3.98 | 0.526 | 0.67 | 0.33;1.33 | 0.260 | 0.49 | 0.19;1.11 | 0.101 |

<sup>a</sup> reference: female, <sup>b</sup> reference: no migration background, <sup>c</sup> reference: German language region, <sup>d</sup> reference: primary education, <sup>e</sup> reference: no radiotherapy; <sup>f</sup> reference: no HSCT, <sup>g</sup> due to the lower n in this analysis, we combined primary and secondary education into in category for children; for adolescents and adults, we kept it separate, HSCT = hematopoietic stem cell transplantation

**Supplementary Table 8.** Sensitivity analysis: Factors associated with sun protection, sunburn, and physician skin examinations in multivariable logistic regression in child, adolescents, and adults CCS, including year of study instead of birth year as exposure

|  | Children (5-15 years) |  |  | Adolescents (16-19 years) |  |  | Adults (≥20 years) |  |  |
| --- | --- | --- | --- | --- | --- | --- | --- | --- | --- |
|  | OR | 95%CI | p | OR | 95%CI | p | OR | 95%CI | p |
| **Sun protection** |  |  |  |  |  |  |  |  |  |
|  | **N = 1,013** |  |  | **N = 476** |  |  | **N = 1,626** |  |  |
| Age at study | 0.78 | 0.71;0.85 | <0.001 | 0.95 | 0.80;1.13 | 0.558 | 1.04 | 1.02;1.07 | <0.001 |
| Male<sup>a</sup> | 0.72 | 0.47;1.09 | 0.125 | 0.37 | 0.24;0.56 | <0.001 | 0.51 | 0.40;0.65 | <0.001 |
| Migration background<sup>b</sup> | 0.63 | 0.38;1.05 | 0.069 | 0.58 | 0.34;0.98 | 0.043 | 0.58 | 0.43;0.80 | 0.001 |
| French/Italian language region<sup>c</sup> | 1.28 | 0.82;2.04 | 0.293 | 1.19 | 0.75;1.90 | 0.461 | 1.01 | 0.77;1.33 | 0.947 |
| Parental education<sup>d</sup> |  |  |  |  |  |  |  |  |  |
| Secondary | 1.96 | 0.97;3.89 | 0.057 | 1.25 | 0.53;2.95 | 0.602 | 1.30 | 0.87;1.93 | 0.197 |
| Tertiary | 1.95 | 0.97;3.79 | 0.054 | 1.71 | 0.69;4.22 | 0.243 | 1.55 | 1.01;2.36 | 0.042 |
| Radiotherapy<sup>e</sup> | 1.33 | 0.77;2.43 | 0.326 | 1.53 | 0.97;2.45 | 0.073 | 1.21 | 0.94;1.56 | 0.149 |
| HSCT<sup>f</sup> | 1.10 | 0.50;2.78 | 0.827 | 1.56 | 0.65;4.08 | 0.333 | 1.02 | 0.61;1.79 | 0.931 |
| Year of study<sup>g</sup> |  |  |  |  |  |  |  |  |  |
| 2010-2013 |  |  |  | 0.44 | 0.28;0.68 | <0.001 | 0.84 | 0.61;1.16 | 0.288 |
| 2015-2017 | 0.96 | 0.58;1.63 | 0.892 |  |  |  |  |  |  |
| 2021-2022 | 0.55 | 0.33;0.92 | 0.021 | 0.49 | 0.26;0.91 | 0.023 | 0.60 | 0.43;0.84 | 0.003 |
| **Sunburn** |  |  |  |  |  |  |  |  |  |
|  | **N = 1,013** |  |  | **N = 542** |  |  | **N = 1,837** |  |  |
| Age at study | 1.12 | 1.05;1.19 | <0.001 | 1.01 | 0.87;1.17 | 0.883 | 0.95 | 0.94;0.96 | <0.001 |
| Male<sup>a</sup> | 0.98 | 0.73;1.34 | 0.920 | 0.72 | 0.51;1.02 | 0.067 | 1.04 | 0.86;1.26 | 0.697 |
| Migration background<sup>b</sup> | 0.54 | 0.35;0.82 | 0.005 | 0.71 | 0.44;1.15 | 0.172 | 0.46 | 0.35;0.62 | <0.001 |
| French/Italian language region<sup>c</sup> | 0.93 | 0.66;1.28 | 0.647 | 1.13 | 0.76;1.66 | 0.551 | 0.87 | 0.70;1.09 | 0.221 |
| Parental education<sup>d</sup> |  |  |  |  |  |  |  |  |  |
| Secondary | 1.09 | 0.58;2.13 | 0.804 | 1.08 | 0.48;2.47 | 0.853 | 1.18 | 0.82;1.70 | 0.376 |
| Tertiary | 0.90 | 0.49;1.76 | 0.759 | 1.33 | 0.58;3.14 | 0.504 | 1.49 | 1.03;2.17 | 0.037 |
| Radiotherapy<sup>e</sup> | 0.67 | 0.43;1.02 | 0.067 | 0.63 | 0.43;0.93 | 0.021 | 0.75 | 0.61;0.92 | 0.006 |
| HSCT<sup>f</sup> | 0.61 | 0.30;1.14 | 0.138 | 1.47 | 0.73;3.00 | 0.286 | 0.62 | 0.39;0.96 | 0.035 |
| Year of study<sup>g</sup> |  |  |  |  |  |  |  |  |  |
| 2010-2013 |  |  |  | 1.03 | 0.68;1.54 | 0.903 | 1.12 | 0.86;1.47 | 0.397 |
| 2015-2017 | 1.57 | 1.07;2.31 | 0.021 | 1.05 | 0.58;1.87 | 0.879 | 1.34 | 0.99;1.82 | 0.060 |
| 2021-2022 | 3.73 | 2.57;5.47 | <0.001 | 1.80 | 1.00;3.31 | 0.052 | 2.57 | 1.89;3.52 | <0.001 |
| **Physician skin examinations** |  |  |  |  |  |  |  |  |  |
|  | **N = 275<sup>i</sup>** |  |  | **N = 538** |  |  | **N = 1,807** |  |  |
| Age at study | 0.86 | 0.73;1.00 | 0.057 | 1.02 | 0.83;1.26 | 0.841 | 1.02 | 1.00;1.04 | 0.020 |
| Male<sup>a</sup> | 0.98 | 0.46;2.11 | 0.958 | 1.38 | 0.85;2.27 | 0.199 | 0.65 | 0.51;0.84 | 0.001 |
| Migration background<sup>b</sup> | 0.83 | 0.32;1.98 | 0.690 | 1.27 | 0.66;2.34 | 0.451 | 1.38 | 0.98;1.94 | 0.064 |
| French/Italian language region<sup>c</sup> | 2.40 | 1.09;5.32 | 0.029 | 1.12 | 0.65;1.89 | 0.674 | 1.38 | 1.04;1.81 | 0.022 |
| Parental education<sup>g</sup> |  |  |  |  |  |  |  |  |  |
| Secondary |  |  |  | 1.50 | 0.50;5.66 | 0.503 | 0.96 | 0.62;1.50 | 0.848 |
| Tertiary | 3.55 | 1.32;11.61 | 0.020 | 1.53 | 0.49;5.92 | 0.497 | 1.27 | 0.81;2.01 | 0.306 |
| Radiotherapy<sup>e</sup> | 2.42 | 0.87;6.27 | 0.076 | 1.14 | 0.67;1.91 | 0.620 | 1.02 | 0.78;1.33 | 0.880 |
| HSCT<sup>f</sup> | 3.82 | 1.42;9.93 | 0.006 | 2.09 | 0.88;4.63 | 0.078 | 2.05 | 1.27;3.23 | 0.003 |
| Year of study |  |  |  |  |  |  |  |  |  |
| 2010-2013 |  |  |  | 0.82 | 0.46;1.46 | 0.511 | 1.31 | 0.93;1.83 | 0.123 |
| 2015-2017 |  |  |  | 0.86 | 0.36;1.91 | 0.721 | 1.23 | 0.82;1.81 | 0.303 |
| 2021-2022 |  |  |  | 1.05 | 0.46;2.24 | 0.909 | 1.03 | 0.67;1.54 | 0.887 |

<sup>a</sup> reference: female, <sup>b</sup> reference: no migration background, <sup>c</sup> reference: German language region, <sup>d</sup> reference: primary education, <sup>e</sup> reference: no radiotherapy; <sup>f</sup> reference: no HSCT; <sup>g</sup> year of study reference category for children is 2007-2013 and for adolescents and adults 2007 to 2009; <sup>h</sup> due to the lower n in this analysis, we combined primary and secondary education into in category for children; for adolescents and adults, we kept it separate; <sup>i</sup> Since we only asked preventive skin examinations for children in 2021/2022, we did not include it study year as exposure for preventive skin examinations in this age group, HSCT = hematopoietic stem cell transplantation

